# Dissected subgroups predict the risk of recurrence of stage II colorectal cancer and select rational treatment

**DOI:** 10.1101/2022.06.20.22276657

**Authors:** Fulong Wang, Shixun Lu, Xin Zhou, Xiaotang Di, Rujia Wu, Gong Chen, Sun Tian

## Abstract

**Background:** Stage II colorectal cancer(CRC) patients after surgery alone have a five-year survival rate of ∼60-80%; the incremental benefit of adjuvant chemotherapy is <5%. Predicting risk of recurrence and selecting effective personalized adjuvant drugs for stage II CRC using formalin-fixed, paraffin-embedded(FFPE) samples is a major challenge.

**Methods:** 1319 stage II CRC patients who enrolled in 2011-2019 at Sun Yat-sen University Cancer Center were screened. RNAseq data of FFPE tumor samples of 222 stage II MSS CRC patients(n.recurrence=47, n.norecurrence=175, median follow-up=41 months) were used to develop a method TFunctionalProg for dissecting heterogeneous subgroups of recurrence, predicting risk of recurrence and proposing adjuvant drugs.

**Results:** TFunctionalProg showed significant predictive values in 222 stage II MSS CRCs. The TFunctionalProg low-risk group had significantly better RFS (validation set (HR=4.78, p-value=1e-4, low-risk group three-year RFS=92.6%, high-risk group three-year RFS=59.7%). TFunctionalProg dissected two subgroups of transition states of stage II MSS CRCs at a high risk of recurrence; each state displays distinct levels of hybrid epithelial-mesenchymal traits, cytotoxic cell inhibition and FOLFOX resistance. Based on mechanisms in two subgroups, TFunctionalProg proposed personalized rational adjuvant drug combinations of immunotherapy, chemotherapy and repurposed CNS drugs. The complementary utility of TFunctionalProg and ctDNA-based prognostic biomarkers were presented.

**Conclusion:** TFunctionalProg was validated using FFPE samples to predict the risk of recurrence and propose rational adjuvant drug combinations for stage II CRCs.

## Introduction

Approximately one third of new cases of colorectal cancer(CRC) are diagnosed with stage II disease. Patients with stage II CRC after surgical resection alone have a five-year survival rate of about 60-80%.[1] The incremental benefit of adjuvant chemotherapy for stage II CRC is less than 5%.[2] One major challenge is the development of molecular diagnostics that can utilize formalin-fixed, paraffin-embedded(FFPE) samples to predict the risk of recurrence of stage II CRC and select effective personalized adjuvant drugs for stage II CRC patients who have a high risk of recurrence. Yet to date, reported successful development of prognostic methods of stage II CRC in FFPE samples are limited. Moreover, current prognostic methods of stage II CRC tend to adopt a simple assumption that all stage II CRC patients with recurrence are a single homogenous group. This overly-simplified disease model does not capture the heterogeneous nature of mechanisms that contribute to recurrence of stage II CRC, thus can not sensitively capture the risk of recurrence and propose effective personalized adjuvant drugs to treat high-risk stage II CRC patients.

Accumulated evidence has shown that stage II CRC patients with recurrence contain heterogeneous subgroups. The first evidence is CRC molecular subtypes. The tumor cells of the epithelial-like subtype and the mesenchymal-like subtype have almost opposite characters, and the tumor microenvironment of the majority of microsatellite instable subtype (MSI) and microsatellite stable (MSS) subtypes are distinct, yet all subtypes contain CRC that developed a recurrence.[3,4] The second evidence is the observation from our previous studies that, in drug resistance to targeted therapy[5], FOLFOX chemotherapy[6] and immunotherapy in CRC[7], the existence of heterogeneous subgroups with distinct gene expression patterns is a repeated phenotype.

Therefore, if the evidence indicates that high-risk stage II CRCs contain heterogeneous subgroups, several questions remain: What are these subgroups? How do the function of these subgroups contribute to recurrence? Should different subgroups of high-risk stage II CRC patients be treated with different adjuvant drugs? What types of signals of these subgroups are preserved in FFPE samples, and how do we find them?

In this report, the clinical data of 1319 stage II Chinese CRC patients enrolled at Sun Yat-sen University Cancer Center in 2011-2019 were screened; the FFPE samples of 222 stage II MSS patients with complete follow-up and treatment data were used to generate RNAseq data. We then developed and validated a novel method TFunctionalProg to dissect subgroups of two different transition states of recurrence and to predict the risk of recurrence of stage II CRC. Based on distinct tumor characters and tumor microenvironment of two different transition states of high-risk stage II CRC, TFunctionalProg proposes personalized rational adjuvant drug combinations of immunotherapy, chemotherapy and repurposed CNS drugs.

## Materials and Methods

Clinical data and follow-up data from 1319 stage II colorectal cancer patients who enrolled in 2011-2019 at Sun Yat-sen University Cancer Center were screened. The main endpoint was recurrence-free survival(RFS). Time to recurrence was calculated from the date of surgery to the date of recurrence confirmed by radiological assessment. Patients who developed a recurrence were defined as the high-risk group. To remove the potential effect of residual disease at the resection margin, patients with time to recurrence less than 2 months were excluded from this study. Patients who did not develop a recurrence were defined as the low-risk group. To remove the potential effect of treatment, patients who received neoadjuvant or adjuvant chemotherapy/radiotherapy were excluded from the low-risk group. The MSI/MSS status of patients was determined by immunohistochemistry for four mismatch repair proteins (*MLH1, MSH2, MSH6, PMS2*). Loss of any of these four proteins was defined as MSI, and the presence of all four proteins was defined as MSS. Because MSI patients have distinct gene expression pattern, and approximately 90% of stage II MSI patients are cured by surgery without any adjuvant chemotherapy, and current guidelines also do not recommend adjuvant chemotherapy for stage II MSI patients, only patients with MSS status were enrolled in this study.[1,8,9] In total, the high-risk group included 47 patients and the low-risk group included 175 patients (n.recurrence=47, n.norecurrence=175, median follow-up=41 months). The patient characteristics are described in Table 1 (**Supplementary Table S1**). Patients or the public were not involved in the design, or conduct, or reporting, or dissemination plans of our research.

RNAseq data of these 222 stage II MSS CRC tumor FFPE samples were used to develop the TFunctionalProg method. Figure 1(**Figure.1**) shows the design of the study and the implementation of the iterative workflow used for training and validating TFunctionalProg. The lab procedure and quality control of RNAseq data, detailed statistical analysis, functional analysis, read out of FOLFOX resistance scores and 14 tumor infiltrating immune cells are described in the eMethod(**Supplementary eMethod**).

**Figure 1.**
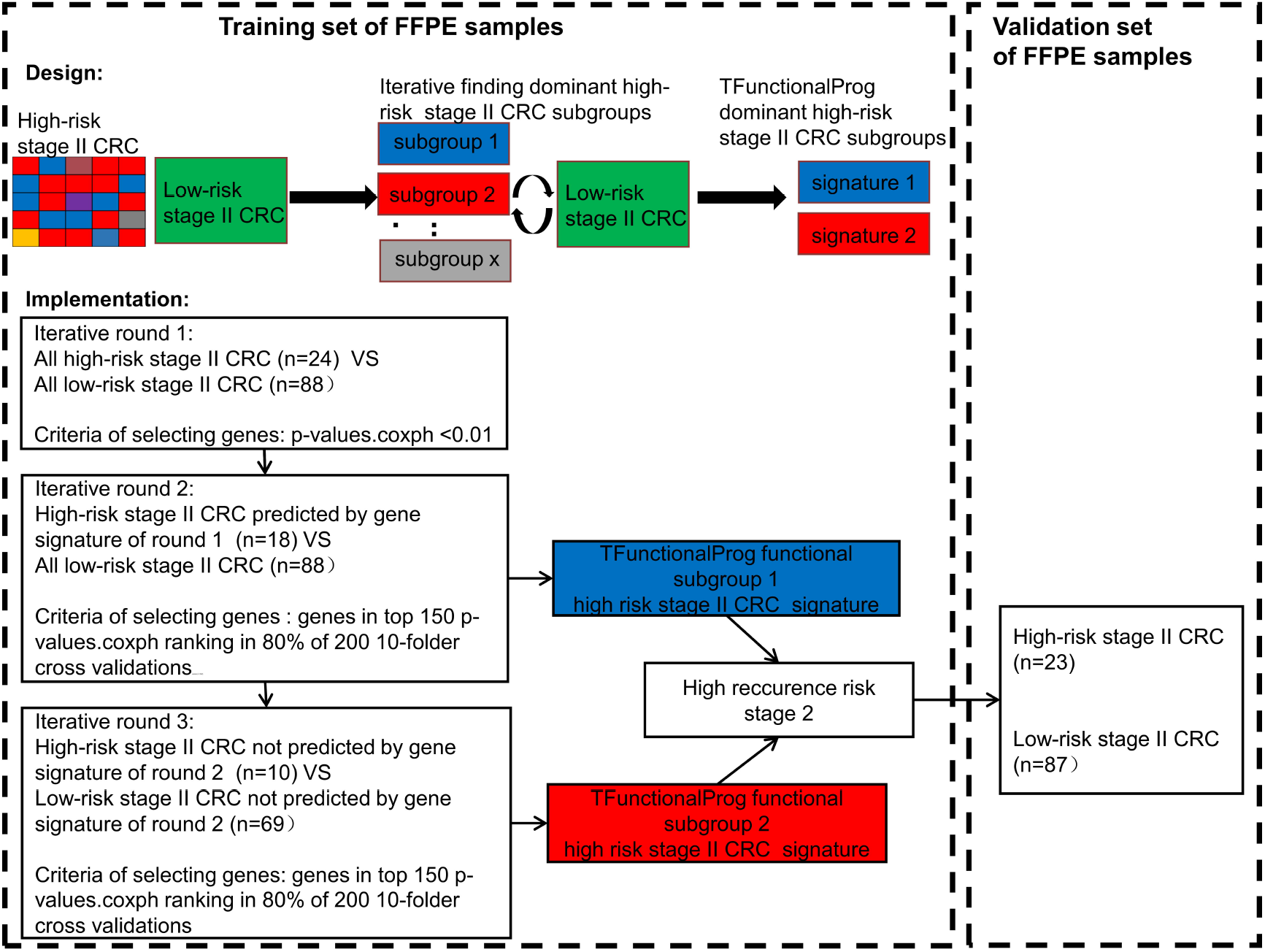
The implementation of the TFunctionalProg method dissects stage II colorectal cancer patients with recurrence into two subgroups, and then analyzes the mechanism of recurrence of the two subgroups separately.

## Results

### Prognostic value of the TFunctionalProg model in stage II CRC tumors

The final prediction method TFunctionalProg consists of two gene sets of the iterative functional prognostic model(n.set1=41 **Supplementary Table S2**, n.set2=40 **Supplementary Table S3**). In the training set of 112 stage II MSS CRCs (24 high-risk and 88 low-risk), the survival analysis resulted in a significant hazard ratio and p-value (p-value<1e-4). The TFunctionalProg high-risk group contained 45.5% of samples. 100% of stage II CRC patients in the TFunctionalProg low-risk group did not develop recurrence within the follow-up period. The TFunctionalProg low-risk group had a three-year RFS rate of 100% and the TFunctionalProg high-risk group had a three-year RFS rate of 58.1% [95% CI, 45.9%-73.5%].

In the independent validation set of 110 stage II MSS CRCs (23 high-risk and 87 low-risk), the hazard ratio and p-value of the survival analysis of the TFunctionalProg method remained significant (HR=4.78, p-value=1e-4). The TFunctionalProg high-risk group contained 38.2% of samples; 89.7% of stage II CRC patients in the TFunctionalProg low-risk group did not develop a recurrence within the follow-up period. The low-risk group had a three-year RFS rate of 92.6% [95% CI, 86.6%-99.1%] and the high-risk group had a three-year RFS rate of 59.7% [95% CI, 46.1%-77.2%] (**Figure 2**).

**Figure 2.**
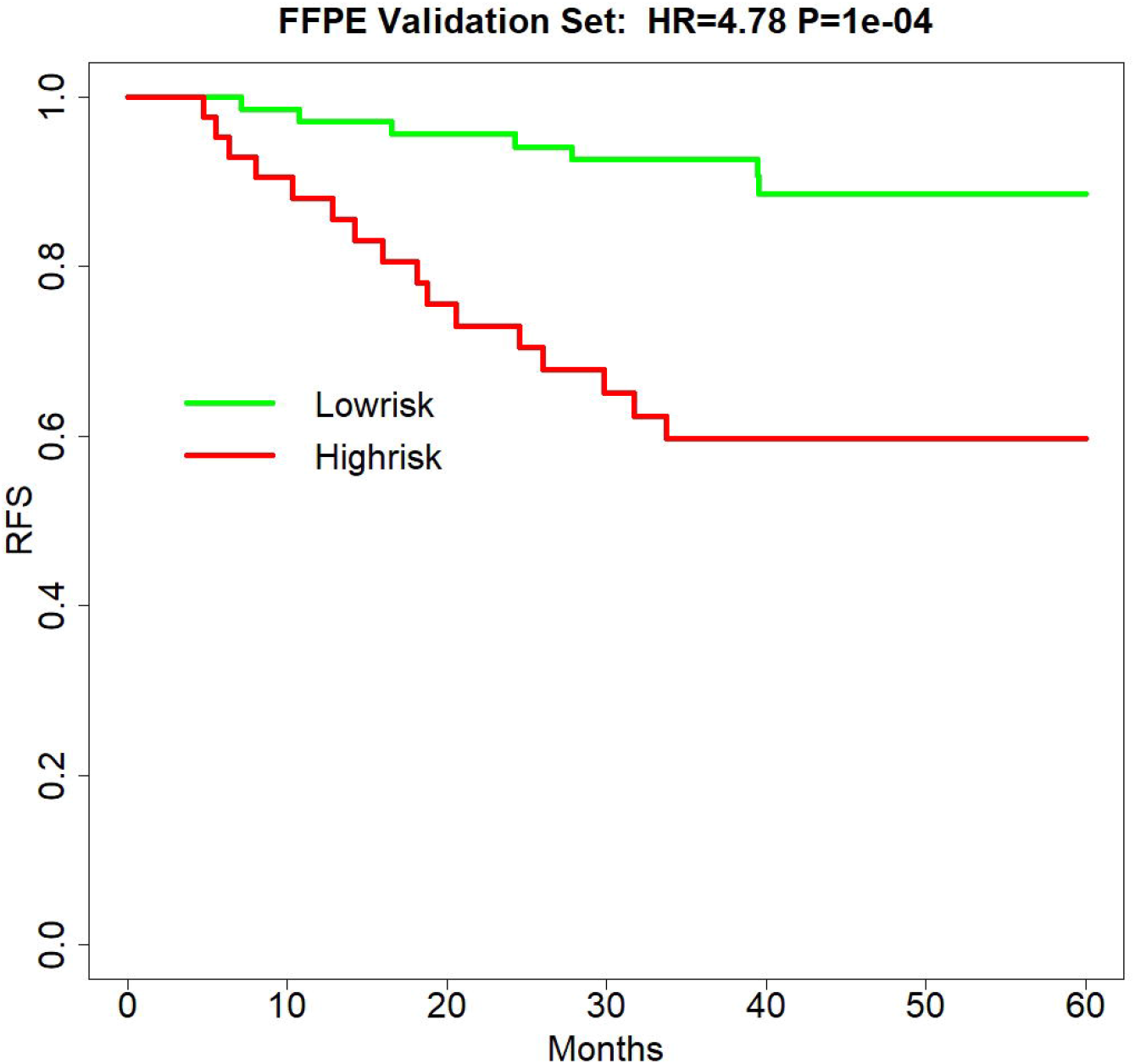
Survival curve of recurrence-free survival in the independent validation set of FFPE samples (HR=4.78, p-value=1e-4). Green is the TFunctionalProg low-risk group and red is the TFunctionalProg high-risk group. The low-risk group had a three-year recurrence-free survival rate of 92.6% and may consider forgoing adjuvant therapy. The high-risk group had a three-year recurrence-free survival rate of 59.7% and contains two subgroups. Based on the different tumor characters of two TFunctionalProg high-risk subgroups, personalized rational adjuvant chemotherapy+drug combinations may be selected.

### TFunctionalProg identified different transition states of metastasis in stage II CRC with implications on the chemotherapy response

The TFunctionalProg model identified two subgroups, both associated with a high risk of recurrence of stage II CRC. TFunctionalProg subgroup 1 (n.set1=41) was characterized by the downregulation of genes involved in the mitotic cell cycle *(PCNA, CDK4, RPA2, MCM6, MCM2*). In addition, ∼50% of the genes in subgroup 1 have a known function in the nucleus (*PCNA, HIST1H2AH, CAV2, SETD1A, HIST1H2BK, RPA2, TNFAIP3, CACYBP, ENO1, HIF3A, UTP18, DUSP23, WT1, CDK4, SNAPC4, PIR, MCM6, MTA2, MCM2, NR2C2AP, E2F8*). The log2 scale of the mean fold changes (high-risk to low-risk) of downregulation was -0.82. This low proliferation index may reflect the characteristics of tumor cells that have already undergone epithelial-mesenchymal transition (EMT). Epithelial markers (*EPCAM, CDH1, EGFR, MET*) were lost in subgroup 1. Interestingly, except for the upregulation of FLT1, most classic mesenchymal markers were not found to be upregulated in subgroup 1 (**blue box, Figure 3, Supplementary Figure S1-S2**). The markers of 14 common tumor infiltrating immune cell types derived from TCGA data showed that tumors in subgroup 1 harbored considerably lower levels of markers of neutrophils (*FCGR3A*), macrophages (*CD68*), T cells (*CD3D, CD3E*), exhausted CD8+ T cells (*CD244, LAG3*), DCs (*HSD11B1*), B cells (*CD19*), CD8+ T cells (*CD8A*) and cytotoxic cells (*GZMA, GZMB, KLRD1, PRF1*) (**blue box, Figure 3, Supplementary Figure S3-S5**).[10] These results suggest that tumors in subgroup 1 tend to lack tumor infiltrating immune cells. Taken together, subgroup 1 may reflect tumor cells that have already initiated EMT, maintain a low proliferation rate and begin to form a cytotoxic cell desert.

**Figure 3.**
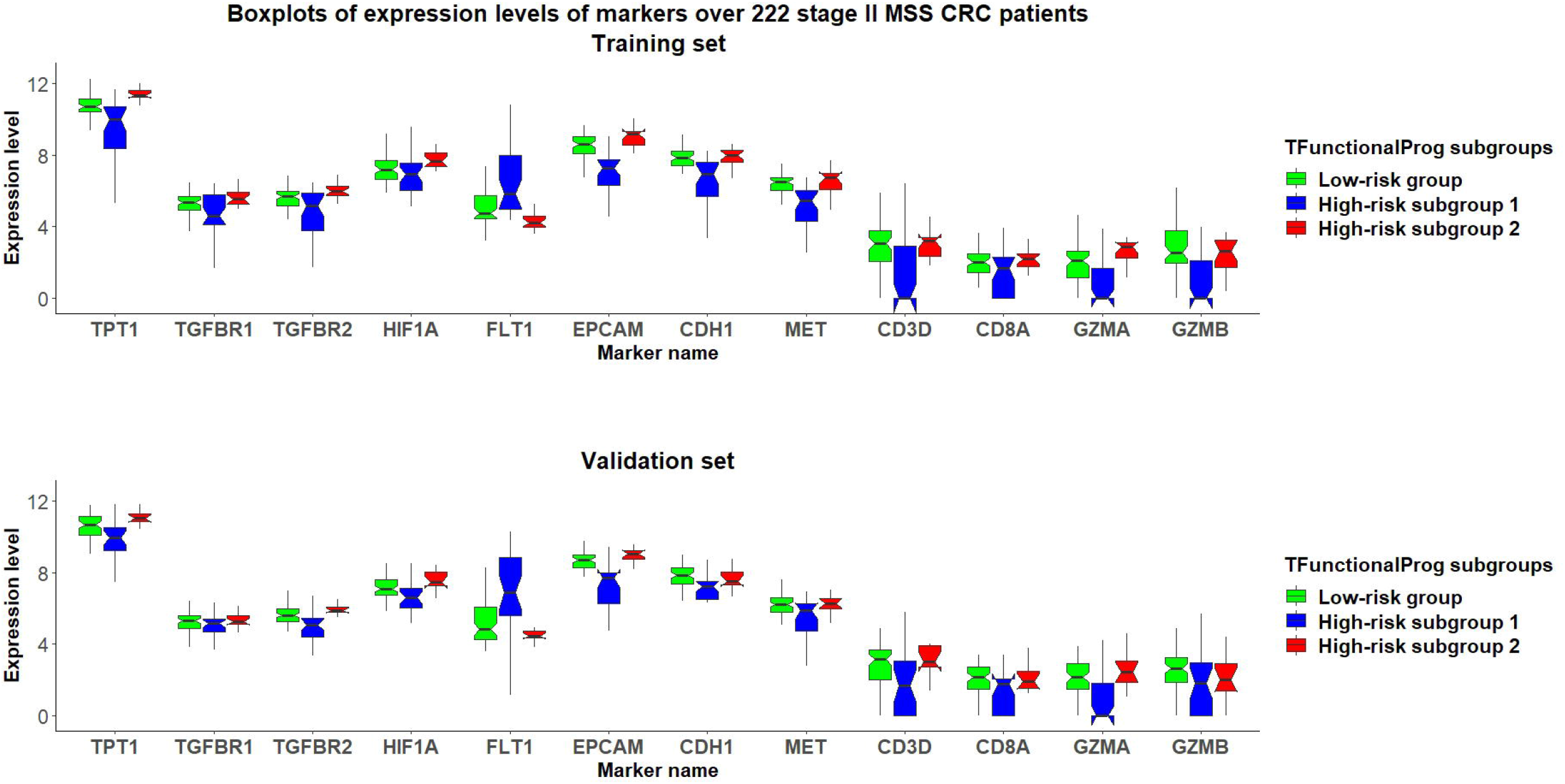
Grouped boxplots of expression levels (Y-axis) of selected markers of epithelial mesenchymal transition (*TPT1, TGFBR1, TGFBR2, FLT1*), hypoxia (*HIF1A*), epithelial phenotype (*EPCAM, CDH1, MET*) and tumor infiltrating cytotoxic T cells (*CD3D, CD8A, GZMA, GZMB*) in 222 stage II MSS CRC patients divided into three subgroups: (1) TFunctionalProg low-risk group (green), (2) TFunctionalProg high-risk subgroup 1 (blue) and (3) TFunctionalProg high-risk subgroup 2 (red). The upper panel is the training set and the lower panel is the independent validation set. The tumor characters and tumor microenvironments of the two TFunctionalProg high-risk subgroups are clearly different and suggest that recurrence of stage II CRC requires transitions from multiple distinct states.

The TFunctionalProg subgroup 2 (n.set2=40) showed comparable counts of tumor infiltrating immune cells, and the level of epithelial markers was maintained (**red box, Figure 3, Supplementary Figure S2-S5**). A tendency towards the upregulation of several known initiators of EMT was observed: *TGFBR1, TGFBR2, TGFB3* and *TPT1* (**red box, Figure 3, Supplementary Figure S1**). Furthermore, the most significant enriched pathways in the signature of subgroup 2 could be mapped to TGF-β receptor signaling and the regulation of cytoplasmic and nuclear SMAD2/3 signaling (percentage=35.3%, p.hypergeometric=9.72e-5, **Supplementary Table S4**). Activating TGF-β pathway is an extensively documented factor of metastasis initiation in CRC.[11–13] Our results are consistent with the known roles of TGF-β; however, our results indicate two additional facts: (1) only ∼35% of the signals of subgroup 2 can be explained by TGF-β and the SMAD2/3 pathway, so activating TGF-β is unlikely the only signal; (2) the high metastatic potential of CRC can be best induced when TGF-β activation is mixed in tumor cells that still maintain some epithelial characters (epithelial markers, **red box, Figure 3, Supplementary Figure S2**). In this state, tumor cells may maintain both epithelial and mesenchymal characters. These results are consistent with the in vivo observation that tumor cells with a hybrid epithelial and mesenchymal phenotype can efficiently reach the circulation and form metastases.[14] Except for activating TGF-β, another dominant character of subgroup 2 is the upregulation of *TPT1*. Loss of *TPT1* results in loss of malignancy of tumor cells.[15] In addition, both the TGF-β pathway and *TPT1* are known factors that suppress anti-tumor immunity. Activation of the TGF-β pathway can initiate the immune exclusion phenotype and TPT1 upregulation can induce myeloid-derived suppressor cells.[16,17] Thus, although the numbers of cytotoxic cells in subgroup 2 were not significantly decreased (**red box, Figure 3, Supplementary Figure S3-S4**), their function may be suppressed. Taken together, subgroup 2, associated with a high risk of recurrence of stage II CRC, likely reflects a state of the initiation of EMT and a hybrid state of epithelial and mesenchymal phenotype. Major drivers include TGF-β and *TPT1* signaling.

The functions of these two TFunctionalProg subgroups suggest that the recurrence of stage II CRC might not be as simple as a binary 0-to-1 transition that directly transits from tumor characters of no recurrence to tumor characters of recurrence. Rather, the development of stage II CRC recurrence might occur through multiple transition states. In our study, there were two stable states in CRC identified: (1) TFunctionalProg subgroup 1 might represent a late stage of EMT where tumor cells have lost almost all epithelial characters (**blue box, Figure 3, Supplementary Figure S2**) and an immune desert has formed (**blue box, Figure 3, Supplementary Figure S3-S5)**. (2) TFunctionalProg subgroup 2 might represent an early stage of EMT where epithelial characters are still maintained by tumor cells, but the signal of EMT initiation is apparent. As a consequence, tumors in subgroup 2 show the upregulation of epithelial characters and the upregulation of some mesenchymal characters (**red box, Figure 3, Supplementary Figure S1-S2**). Our gene expression data in stage II CRC patients agree with the in vivo observation that EMT is a multi-state transition process and proceeds through intermediate transition states, in which hybrid states of epithelial characters and mesenchymal characters exist and form invasive tumors.[14,18,19] The two transition states pose the question as to what factors drive stage II CRC to transit from the early state to the late state of recurrence. This driving factor of the transition between these two states is unclear. Interestingly, *FLT1* was upregulated in TFunctionalProg subgroup 1 (**blue box, Figure 3, Supplementary Figure S1**) and *HIF1A* was upregulated in TFunctionalProg subgroup 2 (**red box, Figure 3, Supplementary Figure S6**); these characters suggest that one driving factor might be hypoxia.[20]

The two TFunctionalProg subgroups also showed different responses to chemotherapy. FOLFOX resistance scores were read out on all samples using the FOLFOX response signature.[6] The results show that CRC patients in TFunctionalProg subgroup 2 had higher FOLFOX resistance scores (p.training=0.034, p.validation=0.001, **red box, panel 1, Figure 4**). These results partially explain why adjuvant chemotherapy has less than a 5% incremental benefit for stage II CRC patients.[2] Subgroup 2 high-risk patients tend to resist FOLFOX, while the tumor cells of subgroup 1 high-risk patients are already in a late stage of EMT and their cytotoxic cells are depleted.

**Figure 4.**
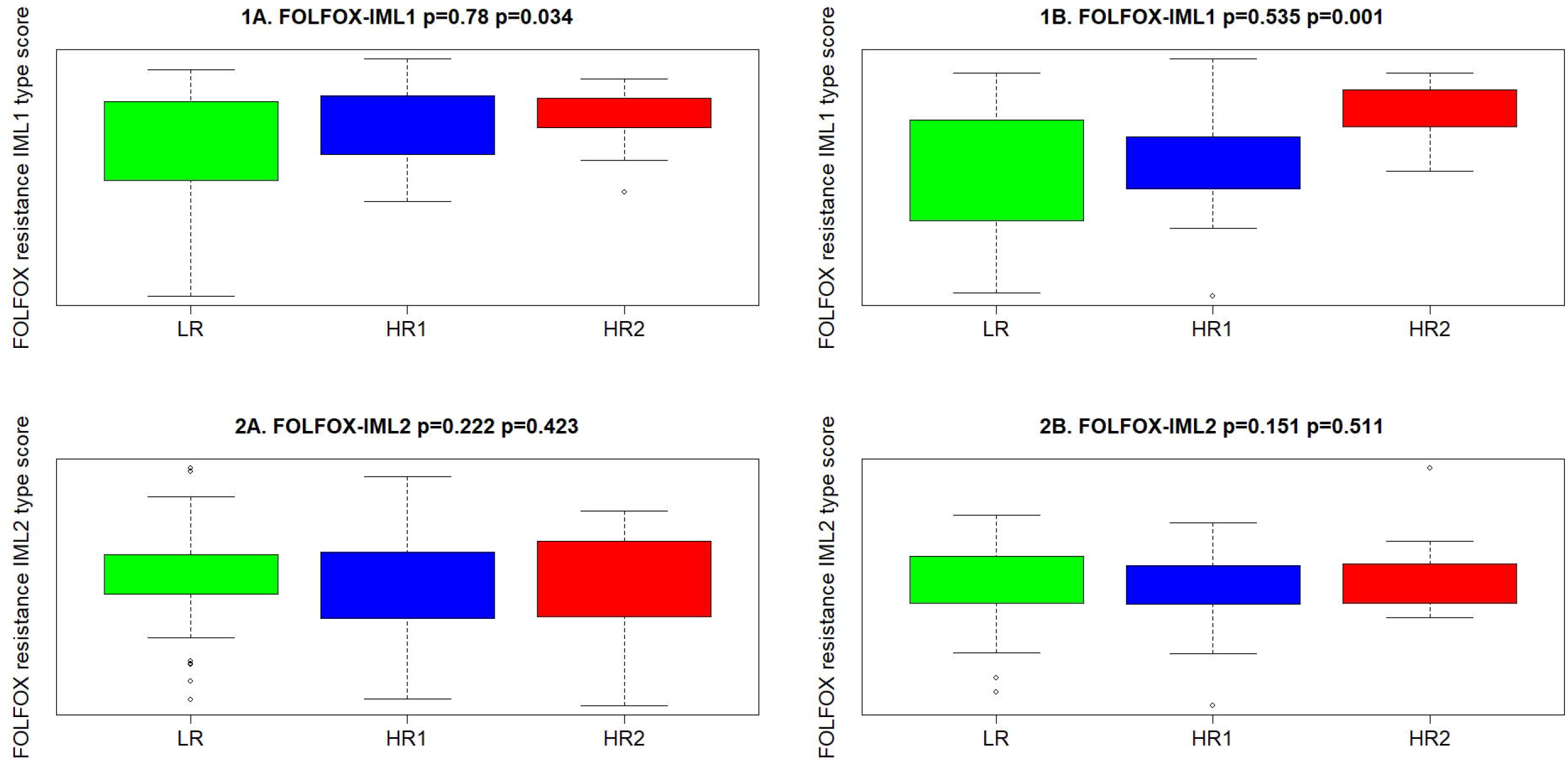
FOLFOX chemotherapy resistance scores of the TFunctionalProg low-risk group (green, LR), TFunctionalProg high-risk subgroup 1 (blue, HR1) and TFunctionalProg high-risk subgroup 2 (red, HR2). Type 1 FOLFOX resistance scores are plotted in panel 1A (training set) and panel 1B (validation set), and type 2 FOLFOX resistance scores are plotted in panel 2A (training set) and panel 2B (validation set). The first p-value is the comparison between the FOLFOX resistance scores of stage II CRC patients in the TFunctionalProg high-risk subgroup 1 (blue, HR1) and the FOLFOX resistance scores of the other stage II CRC patients. The second p-value is the comparison of the FOLFOX resistance scores of stage II CRC patients in the TFunctionalProg high-risk subgroup 2 (red, HR1) compared to the scores of the other stage II colorectal patients. Patients in the TFunctionalProg high-risk subgroup 2 (HR2) showed consistently high type 1 FOLFOX resistance scores in both the training set and validation set (p.training=0.034, p.validation=0.001). The resistance to chemotherapy in the subgroups of high-risk stage II CRC explains why adjuvant chemotherapy only showed a minimal incremental benefit in stage II CRC.

## Discussion

The design of prognostic prediction methods needs to reflect the fundamental logic of biology. High-risk stage II CRC is a heterogeneous group, and the recurrence of stage II CRC is likely a multi-state transition. The underlying design of a prediction method needs to identify these distinct subgroups to capture the biology of the risk of recurrence of stage II CRC. TFunctionalProg was validated on FFPE samples and can be implemented as a diagnostic tool to identify stage II CRC patients at a high risk of recurrence.

The TFunctionalProg low-risk group of stage II CRC has a three-year RFSrate of >92% and may consider forgoing adjuvant therapy. The TFunctionalProg high-risk group of stage II CRC has a three-year RFS rate of <60%. The current observed incremental benefit of adjuvant chemotherapy for stage II CRC patients is <5%.[2] Biomarkers are already used to select stage III CRC patient subpopulations to receive personalized adjuvant chemotherapy+immunotherapy.[21,22] It would be interesting to explore further whether different tumor characters of TFunctionalProg high-risk subgroup 1 and TFunctionalProg high-risk subgroup 2 could help select different personalized rational chemotherapy+drug combinations to increase the benefit of adjuvant treatment for stage II CRC patients.[2] Three drug combinations are proposed for further investigation (**Figure 5**):

**Figure 5.**
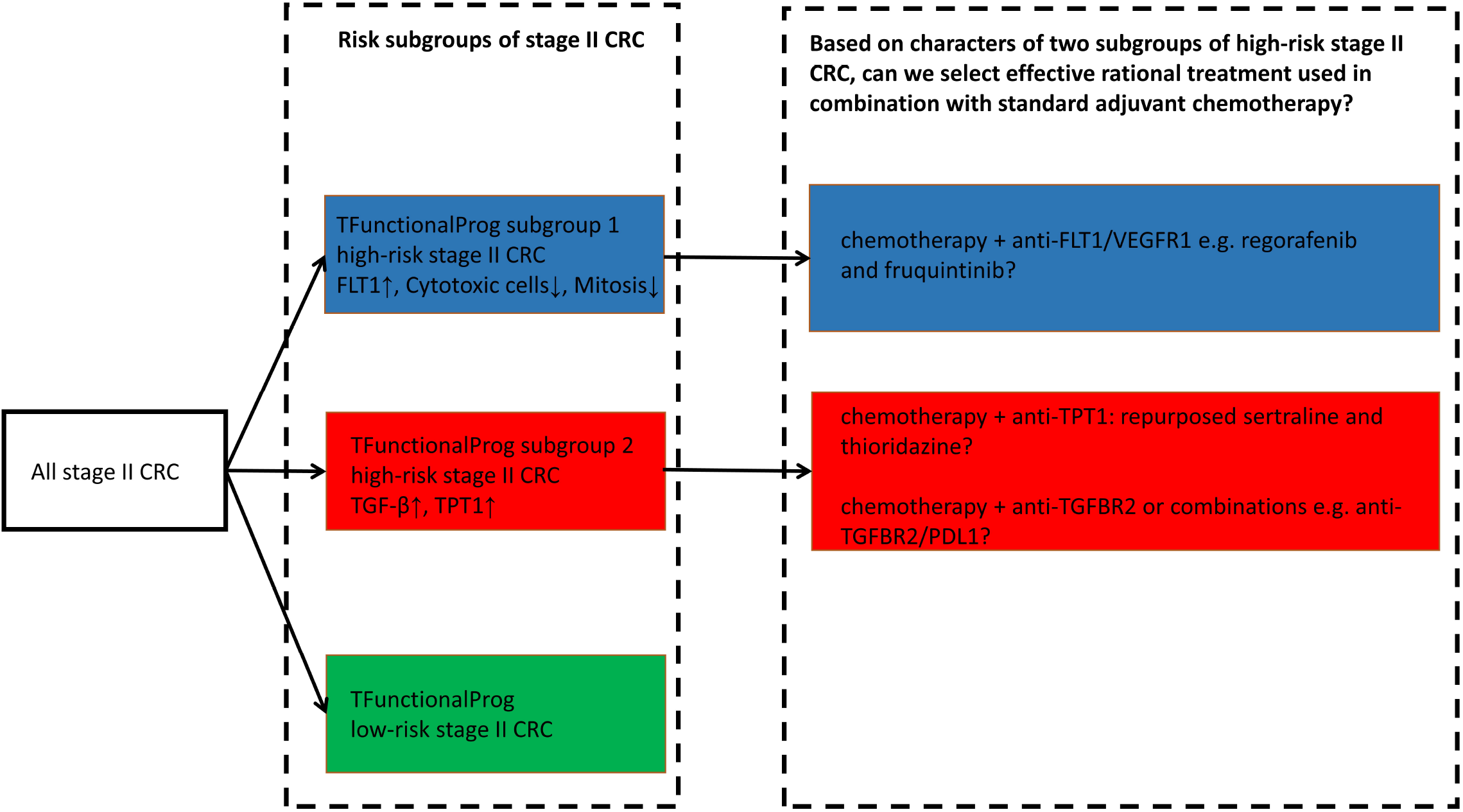
TFunctionalProg subgroups of the risk of recurrence of stage II CRC: low-risk group (green, LR), high-risk subgroup 1 (blue, HR1) and high-risk subgroup 2 (red, HR2). Different tumor characters and tumor microenvironments in TFunctionalProg high-risk subgroup 1 and TFunctionalProg high-risk subgroup 2 suggest that standard chemotherapy may be combined with different rational personalized treatment options to increase the benefit of adjuvant treatment in stage II CRC. This scheme proposes three drug combinations for further investigation.

1. TFunctionalProg high-risk subgroup 2 stage II CRC patients show a dependence on TGF-β pathway activation and *TPT1* (**red box, Figure 3**). It is important to note that the number of CD8+ T cells and cytotoxic cells is not significantly decreased in subgroup 2. Therefore, TFunctionalProg high-risk subgroup 2 CRC patients might represent a patient subpopulation that could be potentially treated by an anti-*PDL1/TGFBR2* bispecific antibody.[13,23]
2. TFunctionalProg high-risk subgroup 2 stage II CRC patients also showed upregulation of *TPT1* (**red box, Figure 3**). *TPT1/TCTP* is a switch regulating tumor reversion. Two central nervous system (CNS) drugs, sertraline and thioridazine, directly bind to *TPT1/TCTP* and inhibit its function.[24] In human CRC cell lines, sertraline and thioridazine, alone or in combination with 5-FU, showed significant inhibition of tumor growth.[25] A biomarker-driven drug repurposing of CNS drugs to treat TFunctionalProg high-risk subgroup 2 stage II CRC patients may be further explored.
3. TFunctionalProg high-risk subgroup 1 stage II CRC patients showed depletion of CD8+ T cells and cytotoxic cells (**blue box, Figure 3**) and upregulation of *FLT1* (**blue box, Figure 3**). *FLT1/VEGFR1* is a vascular endothelial growth factor receptor and its ligands include *VEGFA, VEGFB* and *PlGF*. The anti-*VEGFA* drug bevacizumab is a standard treatment for metastatic CRC and is associated with a benefit in terms of overall survival. Anti-*FLT1/VEGFR1* drugs that are already used in CRC include aflibercept and tyrosine kinase inhibitors such as regorafenib and fruquintinib. Patients with refractory metastatic CRC treated with regorafenib or fruquintinib showed better overall survival.[26] Whether stage II CRC patients benefit from regorafenib or fruquintinib in the adjuvant setting is unknown, but the character of TFunctionalProg high-risk subgroup 1 stage II CRC patients may represent a rational subpopulation in which to start clinical studies.

A rapidly adapted biomarker for predicting stage II CRC at a high risk of recurrence is postoperative circulating tumor DNA(ctDNA) from blood samples. Postoperative positive ctDNA status is an indicator of minimal residual disease and is associated with a high risk of recurrence.[27] However, in three independent clinical trials including the DYNAMIC trial of 291 stage II CRC patients and a recent multi-center study of 240 stage II&III CRC patients at our cancer center, only 35-41% of recurrences of CRC patients are detected by postoperative ctDNA-based prognostic markers(sensitivity for stage II CRC=35%, **Supplementary Table S5**).[28–30] In all studies, ctDNA positive groups had a significantly higher recurrence risk than ctDNA negative groups. Yet, in all studies, ctDNA failed to detect ∼60% of the stage II CRC patients who developed a recurrence. Thus, at least for current ctDNA-based prognostic methods for stage II CRC patients, ctDNA positive status can be used as a biomarker for adjuvant therapy, but whether ctDNA negative status should be used as the criteria to rule out adjuvant therapy is debatable. Postoperative ctDNA-based biomarkers and gene expression-based biomarkers can complement each other. The sensitivity of TFunctionalProg in predicting the recurrence of stage II CRC is between 70% (independent validation) and 85% (leave one out cross-validation training + independent validation), and TFunctionalProg provides rational selections of therapeutic drugs for stage II CRC patients at a high risk of recurrence. Further, it would also be interesting to investigate whether circulating tumor cells of ctDNA-positive stage II CRCs correlates with a TFunctionalProg high-risk subgroup, and this may help the understanding of the functionality and the origin of heterogenous circulating tumor cells.[31] TFunctionalProg utilize standard FFPE slides that can be stored at room temperature and provides an alternative to ctDNA-based prognostic biomarkers.

To conclude, TFunctionalProg was validated on standard FFPE slides to predict the risk of recurrence of stage II CRC and shows high sensitivity in terms of detecting stage II CRC patients at a high risk of recurrence. Considering high-risk stage II CRC patients, TFunctionalProg dissected two different subgroups of transition states of recurrence and proposed different personalized rational adjuvant drug combinations of chemotherapy, immunotherapy and repurposed CNS drugs.

## Data Availability

RNAseq data of 81 genes used in TFunctioanlProg method will be deposit as online supplementary data.

## Ethics approval and patient consent statement

Medical Ethical Committee of Sun Yat-sen University Cancer Center gave ethical approval for this work(nr. B-0440-1).

## Funding

The authors declare no funding.

## Competing interests statement

RW and ST owns stocks and shares and/or stock options in Carbon Logic Biotech Ltd. ST is an inventor of a patent application related to this work. All remaining authors have declared no conflicts of interest.

## Author contributions

FW: patients, collection and/or assembly of data, data analysis and interpretation, manuscript writing first draft, final approval of manuscript

SL: patients, collection and/or assembly of data, final approval of manuscript

XZ: collection and/or assembly of data, final approval of manuscript

XD: data analysis and interpretation, final approval of manuscript

RW: collection and/or assembly of data, data analysis and interpretation, final approval of manuscript

GC: concept/design, patients, collection and/or assembly of data, final approval of manuscript

ST: concept/design, data analysis and interpretation, manuscript writing first draft, final approval of manuscript

## Supplementary Tables

**Supplementary Table S1**. Main clinicopathological characteristics of formalin-fixed, paraffin-embedded samples of 222 stage II MSS CRC patients.

**Supplementary Table S2**. List of genes in the signature of TFunctionalProg high-risk subgroup 1.

**Supplementary Table S3**. List of genes in the signature of TFunctionalProg high-risk subgroup 2.

**Supplementary Table S4**. Enriched biological pathways of genes in the signature of TFunctionalProg high-risk subgroup 2.

**Supplementary Table S5**. Sensitivity of ctDNA-based prognostic methods for the prediction of the risk of recurrence of stage II CRC.

## Supplementary Figures

**Supplementary Figure S1-S6**. Boxplots of the expression levels of markers of mesenchymal phenotype and epithelial-mesenchymal transition(**Figure S1**), epithelial phenotype(**Figure S2**), 14 common tumor infiltrating immune cells (Treg cells, Th1 cells, T cell, NK cells, NK CD56^dim^ cells neutrophils mast cells macrophages exhausted CD8 cells DCs cytotoxic cells, CD8 T cells, CD45 cells, B cells)(**Figure S3-S5**), and hypoxia(**Figure S6**) in 222 stage II CRC patients divided into three subgroups. (1) TFunctionalProg low-risk group (green, LR), (2) TFunctionalProg high-risk subgroup 1 (blue, HR1) and (3) TFunctionalProg high-risk subgroup 2 (red, HR2). Panel A is the training set and panel B is the independent validation set. The gene names of the markers are shown at the top of the boxplot. The first p-value is the comparison between expression levels of a marker in high-risk subgroup 1 (blue, HR1) and expression levels of this marker in the other groups. The second p-value is the comparison between expression levels of a marker in high-risk subgroup 2 (red, HR2) and expression levels of this marker in the other groups.

## Notes

### Author Declarations

Medical Ethical Committee of Sun Yat-sen University Cancer Center gave ethical approval for this work(nr. B-0440-1). Patient consent is waived.

### Summary of Updates

Updated 2 missing values in the supplementary table

