## Supplementary Table for "Dissected subgroups predict the risk of recurrence of stage II colorectal cancer and select rational treatment"

**Supplementary Table S1.** Main clinicopathological characteristics of formalin-fixed, paraffin-embedded samples of 222 stage II MSS CRC patients.

| Clinical Characteristic | Training Set (n = 112) n(%) | Validation Set (n = 110) n(%) |
|---|---|---|
| Sex | | |
| Male | 61(54.5) | 65(59.1) |
| Female | 51(45.5) | 45(40.9) |
| Median age, years | 62 | 61 |
| Median follow-up(months) | 41.3 | 41.2 |
| Localization | | |
| Left | 47(42.0) | 58(52.7) |
| Right | 29(25.9) | 27(24.5) |
| Rectum | 36(32.1) | 25(22.7) |
| T | | |
| 3 | 79(70.5) | 81(73.6) |
| 4 | 33(29.5) | 29(26.4) |
| MSS/MSI | | |
| MSS | 112(100) | 110(100) |
| MSI | 0(0) | 0(0) |
| Recurrence developed during the follow-up period | | |
| No | 88(78.6) | 87(79.1) |
| Yes | 24(21.4) | 23(20.9) |
| Chemotherapy | | |
| No | 97(86.6) | 94(85.5) |
| Yes | 14(12.5) | 15(13.6) |
| Unknown | 1(0.9) | 1(0.9) |

**Supplementary Table S2.** List of genes in the signature of TFunctionalProg high-risk subgroup 1

| **Genename** | **StatisticsOccurenceInCrossvalidation** | **RegulationDir_Reccurence(Highrisk)Group** |
|---|---|---|
| ATG101 | 200 | Down |
| ATP5ME | 169 | Down |
| CACYBP | 177 | Down |
| CAV2 | 190 | Up |
| CCDC137 | 187 | Down |
| CDK4 | 167 | Down |
| CKAP4 | 194 | Down |
| CLPP | 183 | Down |
| COA6 | 178 | Down |
| DCXR | 185 | Down |
| DUSP23 | 200 | Down |
| E2F8 | 194 | Down |
| EIF3I | 178 | Down |
| ELOB | 194 | Down |
| ENO1 | 199 | Down |
| EPHB2 | 200 | Down |
| FARSA | 200 | Down |
| HIF3A | 200 | Up |
| HIST1H2AH | 182 | Down |
| HIST1H2BK | 165 | Down |
| IQANK1 | 172 | Down |
| MCM2 | 200 | Down |
| MCM6 | 192 | Down |
| MTA2 | 198 | Down |
| NR2C2AP | 172 | Down |
| PCNA | 198 | Down |
| PIR | 188 | Down |
| PLIN3 | 194 | Down |
| PRDX2 | 167 | Down |
| ROMO1 | 196 | Down |
| RPA2 | 162 | Down |
| SETD1A | 176 | Down |
| SMPD2 | 197 | Down |
| SNAPC4 | 185 | Down |
| SPATA6 | 197 | Up |
| ST13 | 182 | Down |
| THAP4 | 192 | Down |
| TNFAIP3 | 185 | Down |
| TP73-AS1 | 200 | Up |
| UTP18 | 174 | Down |
| WT1 | 186 | Up |

**Supplementary Table S3.** List of genes in the signature of TFunctionalProg high-risk subgroup 2

| Genename | StatisticsOccurenceInCrossvalidation | RegulationDir_Reccurence(Highrisk)Group |
|---|---|---|
| APLP2 | 181 | Up |
| BTF3 | 198 | Up |
| CAB39 | 177 | Up |
| COPB2 | 176 | Up |
| CRTAP | 200 | Up |
| CSF1 | 200 | Down |
| CSNK1A1 | 168 | Up |
| DSG2 | 188 | Up |
| EIF1AY | 192 | Up |
| ENDOD1 | 200 | Up |
| FCHO2 | 200 | Up |
| FRRS1 | 161 | Up |
| FUT6 | 166 | Down |
| HIST1H2AC | 200 | Up |
| IGFLR1 | 182 | Down |
| ITM2B | 181 | Up |
| MEDAG | 164 | Up |
| MYL12A | 200 | Up |
| MYL12B | 200 | Up |
| OCLN | 191 | Up |
| PAK1 | 169 | Up |
| PFDN5 | 168 | Up |
| PHLDB2 | 163 | Up |
| PMS2P4 | 198 | Up |
| RAB27B | 181 | Up |
| RALB | 200 | Up |
| RALBP1 | 177 | Up |
| RHOA | 194 | Up |
| RSF1 | 180 | Up |
| SEL1L | 195 | Up |
| SPON1 | 188 | Up |
| SPPL3 | 178 | Up |
| TM9SF2 | 184 | Up |
| TMEM30B | 200 | Up |
| TPT1 | 164 | Up |
| TRIM65 | 194 | Down |
| TXNDC17 | 176 | Up |
| VAPA | 196 | Up |
| YWHAE | 200 | Up |
| ZNF74 | 192 | Down |

**Supplementary Table S4.** Enriched biological pathways of genes in the signature of TFunctionalProg high-risk subgroup 2.

| **Biological pathway** | **No. of genes in the dataset** | **Percentage of genes** | **P-value (Hypergeometric test)** |
|---|---|---|---|
| Regulation of cytoplasmic and nuclear SMAD2/3 signaling | 6 | 35,29411765 | 9,72494E-05 |
| TGF-beta receptor signaling | 6 | 35,29411765 | 9,72494E-05 |
| Regulation of nuclear SMAD2/3 signaling | 6 | 35,29411765 | 9,72494E-05 |
| ALK1 signaling events | 6 | 35,29411765 | 0,000129245 |
| ALK1 pathway | 6 | 35,29411765 | 0,000136088 |
| Alpha6Beta4Integrin | 3 | 17,64705882 | 0,000263138 |
| FoxO family signaling | 3 | 17,64705882 | 0,000279831 |
| Apoptotic cleavage of cell adhesion proteins | 2 | 11,76470588 | 0,000446654 |
| Semaphorin interactions | 3 | 17,64705882 | 0,000617129 |
| Regulation of p38-alpha and p38-beta | 4 | 23,52941176 | 0,00081309 |
| EPHA2 forward signaling | 2 | 11,76470588 | 0,001144431 |
| p38 MAPK signaling pathway | 4 | 23,52941176 | 0,001382216 |
| Sema4D induced cell migration and growth-cone collapse | 2 | 11,76470588 | 0,001538632 |
| S1P2 pathway | 2 | 11,76470588 | 0,002151029 |
| CDC42 signaling events | 7 | 41,17647059 | 0,002289032 |
| Sema4D in semaphorin signaling | 2 | 11,76470588 | 0,002319424 |
| PLK1 signaling events | 3 | 17,64705882 | 0,002522717 |
| Regulation of CDC42 activity | 7 | 41,17647059 | 0,002530136 |
| BMP receptor signaling | 4 | 23,52941176 | 0,002670031 |
| Polo-like kinase signaling events in the cell cycle | 3 | 17,64705882 | 0,002884017 |
| IL1-mediated signaling events | 4 | 23,52941176 | 0,003030502 |
| Lissencephaly gene (LIS1) in neuronal migration and development | 2 | 11,76470588 | 0,003053283 |
| Netrin-mediated signaling events | 2 | 11,76470588 | 0,003251668 |
| Signal transduction by L1 | 2 | 11,76470588 | 0,003882157 |
| AP-1 transcription factor network | 6 | 35,29411765 | 0,004317945 |
| p75 NTR receptor-mediated signalling | 2 | 11,76470588 | 0,004804158 |
| Apoptotic cleavage of cellular proteins | 2 | 11,76470588 | 0,004804158 |
| Integrin-linked kinase signaling | 6 | 35,29411765 | 0,005594035 |
| EphrinA-EPHA pathway | 2 | 11,76470588 | 0,005817424 |
| Apoptotic execution phase | 2 | 11,76470588 | 0,007209553 |
| TNF receptor signaling pathway | 4 | 23,52941176 | 0,007283198 |
| Signaling events mediated by PTP1B | 2 | 11,76470588 | 0,008421488 |

**Supplementary Table S5.** Sensitivity of ctDNA-based prognostic methods for the prediction of risk of recurrence of stage II CRC.

| **Reference in the manuscript** | **Number of Patient** | **Patient population** | **Sensitivity of predicting recurrence by postoperative ctDNA** |
|---|---|---|---|
| Chen G, Peng J, Xiao Q, et al. Postoperative circulating tumor DNA as markers of recurrence risk in stages II to III colorectal cancer. *J Hematol OncolJ Hematol Oncol*. 2021;14(1):80. doi:10.1186/s13045-021-01089-z | 240 | Stage II&III colorectal cancer | 38% (12 detected by ctDNA/32 recurrence) |
| Reinert T, Henriksen TV, Christensen E, et al. Analysis of Plasma Cell-Free DNA by Ultradeep Sequencing in Patients With Stages I to III Colorectal Cancer. *JAMA Oncol.* 2019;5(8):1124-1131. doi:10.1001/jamaoncol.2019.0528 | 94 | Stage I-III colorectal cancer | 41% (7 detected by ctDNA/17 recurrence) |
| Tie J, Cohen JD, Lahouel K, *et al.* Circulating Tumor DNA Analysis Guiding Adjuvant Therapy in Stage II Colon Cancer. *N Engl J Med* 2022;**386**:2261–72. doi:10.1056/NEJMoa2200075 | 291 | Stage II | 35% (8 detected by ctDNA/23 recurrence) |
